# Flow-mediated dilatation: learning curve study with a novice operator

**DOI:** 10.1101/2023.09.11.23295348

**Authors:** Arko S. Dhar, Marie Fisk, Carmel M. McEniery, Domonkos Cseh

## Abstract

**Introduction:** Brachial artery flow-mediated dilatation (FMD) is commonly measured to assess endothelial function in experimental and clinical research studies. However, attaining reproducible results can be challenging, especially among inexperienced operators. Until now, no detailed learning curve study of novice operators has been published.

**Methods:** We assessed the learning curve of FMD measurement in one novice operator. Following a one-week basic training period, the operator performed duplicate measurements on 6-8 individuals per week, for six weeks. The operator followed the recommendations of the most recent FMD measurement guideline. Duplex ultrasound was used for the simultaneous and continuous measurement of brachial artery diameter and local blood flow velocity. Following a 1-minute recording of baseline diameter (D), FMD was measured after a 5-minute period of occlusion of the proximal forearm vessels. Inter-session coefficient of variation (CV) values for D and FMD were calculated for each week.

**Results:** The majority of participants did not have any cardiovascular disease history. The number of volunteers assessed each week were the following: 8, 7, 7, 6, 7, 7 individuals, from week 1 to week 6. CV values for both D and FMD exhibited a decreasing trend over the training period, ending at 1.73% for D and 14.24% for FMD at week 6. The CV values at week 6 were within the range outlined in the most recent FMD guideline for proficiency.

**Conclusions:** Within a reasonable timeframe, and with careful adherence to measurement guidelines, the attainment of sound reproducibility in FMD measurements by a novice operator is feasible.

## Introduction

Brachial flow-mediated dilatation (FMD) measurement is a major, non-invasive technique, for the assessment of vascular endothelial function.^1, 2^ FMD is frequently measured in experimental and clinical research studies, especially in the context of vascular treatment responses.^3^ Impaired endothelium-dependent dilatation of the brachial artery provides important information about subclinical organ damage progression in cardiovascular diseases, correlating with pathophysiological markers such as albuminuria and increased pulse wave velocity, even after adjustment for known risk factors.^4^ Low FMD is associated with coronary endothelial dysfunction,^5^ and an independent predictor of in-stent stenosis following single-vessel coronary interventions.^6^ Furthermore, progressively higher FMD values have been associated with reduced risk of cardiovascular events in both low and high-risk populations.^7^ Current guidelines are encouraging the need for experience, based on the attainment of low inter-session coefficient of variation (CV) values.^1^ However, there has not been a published study outlining the content, timeframe, and feasibility of procedural training of completely novice operators. The standardized training regimen presented by Vanoli et al. for the measurement of carotid intima-media thickness, that is another non-invasive, methodologically less complex technique, provides a great example for learning curve studies.^8^ Since the heavy burden of cardiovascular disease on society necessitates that more professionals, especially those without prior experience, become familiar with available non-invasive research techniques for assessing vascular function, we aimed in this study to provide an outline for what a standardized FMD training regimen could entail.

## Methods

The data that support the findings of this study are available from the corresponding author upon reasonable request.

### Training

The training procedure was carried out by a single novice (A.S.D.) with no prior experience in ultrasound measurements. The program began with a 2-hour lecture covering the principles of ultrasonographic imaging and FMD, focusing both on the biophysical and physiological background of the technique. Afterwards, the novice operator was introduced to the portable ultrasound system (MyLab25Gold, Esaote, Genoa, Italy). Subsequently, eight basic scans were carried out by the novice. These scans were meant to give the novice operator familiarity with the basic settings of the ultrasound system and analysis software (Cardiovascular Suite, FMD Studio, Version 4, Quipu, Pisa, Italy). The ability to properly visualize a longitudinal section of the brachial artery with clear vascular boundaries was emphasized. At the end of this period, the operator was able to execute the basic steps of the procedure. After these initial steps, the operator began carrying out FMD measurements on the official subjects of the study. To assess the reproducibility of the measurements, 6-8 individuals were examined each week. Two measurements were performed on each individual on different days, at similar time of the days (+/-1 hour). During the first week, the experienced researcher (D.C.) watched the measurements and during and after each measurement, provided detailed feedback to the novice to help improvement. From the second week, the novice performed the measurements alone, and the recordings were checked and discussed with the experienced researcher afterwards.

### Study population

Volunteers were recruited from the Division of Experimental Medicine and Immunotherapeutics (EMIT) at the University of Cambridge. FMD measurements in the EMIT Division are performed as a part of a larger study (Influence of Age, Weight and Ethnic background on blood pressure: AWE study) that received favorable opinion from the local Research Ethics Committee and our study was performed within the confines of this larger study (ethical permission number: 16/WM/0485). Prior to examination, the subjects were given information about the measurements, and verbal informed consent was received. According to the recent guideline, subjects were asked to fast and refrain from smoking or tobacco consumption for at least 6 hours prior to measurement. They were also asked to refrain from vigorous exercise and alcoholic beverage or stimulant drink consumption for at least 24, and 12 hours, respectively, prior to the measurement.^1^ Anthropometric characteristics of the subjects (height, weight, body mass index), medical history, medications, and smoking status were recorded. Drug withdrawal was not required.

### Flow-mediated dilatation measurement

Subjects were asked to rest in the supine position for 10 minutes in a temperature-controlled (22 ±1°C), dimly lit room. Following this, two automatic blood pressure readings (HEM-907 Digital Blood Pressure Monitor, OMRON Healthcare, Illinois, USA) were taken 1 minute apart. A cuff was then placed over the right forearm distal to the elbow crease, with the arm being at the level of the heart. A small vacuum mattress was used to support and stabilize the upper arm of the subjects, and a probe holder was used to stabilize and maintain the position of the 12 MHz linear array transducer (LA523, Esaote, Genoa, Italy). The measurements were performed using duplex ultrasound. Image depth was 3 cm, the insonation angle for blood flow velocity measurement was 70°, and the same calibration settings were used for each measurement throughout the whole study. The measurement began with a 1-minute period to measure baseline brachial artery diameter (D), followed by a 5-minute occlusion period in which the cuff around the forearm was inflated to 50 mmHg above systolic blood pressure. After this occlusion period, the cuff was fully deflated, and the change in arterial diameter (ΔD) due to reactive hyperemia was measured. FMD was calculated as a percentage according to the following formula: 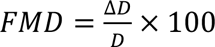. Shear rate (SR) was calculated as follows: 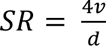, where v represents velocity of blood flow, and d represents diameter. Major output variables of the measurements were the following: D, FMD, baseline SR and maximal SR.

In order to achieve precision with respect to placement of the probe and the cuff, distances from the elbow crease to both the probe and cuff were measured using a removable skin marker and a ruler, and documented. These distances were then used for placement of the probe and the cuff in the follow-up measurements for each subject. Additionally, the angle of probe position was measured using a protractor and documented. This angle was likewise used to orient the ultrasound probe in the subsequent measurement on the same subject.

### Data analysis

In order to properly follow the improvement over the learning period, we opted not to discard any measurements. Statistical analysis was carried out using IBM SPSS v. 28 (IBM Corporation, Somers, NY, USA). Coefficient of variation (CV) was calculated to assess the inter-session reproducibility of the measurements. It was calculated as follows: 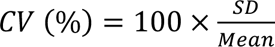, where SD represents within-subject standard deviation. SD was calculated as follows: 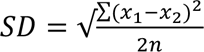, where n is the number of data pairs and x_1_ and x_2_ are the duplicate measurements.^9^ Data are presented as n (%) and, due to small sample sizes and non-normal distributions, medians [interquartile ranges (IQR)].

## Results

Anthropometric, hemodynamic, and relevant demographic data are shown in **Table 1**. The majority of the volunteers did not have any history of cardiovascular disease.

**Table 1.** Subject characteristics.

|  | **Week 1**<br>**n = 8** |  | **Week 2**<br>**n = 7** |  | **Week 3**<br>**n = 7** |  | **Week 4**<br>**n = 6** |  | **Week 5**<br>**n = 7** |  | **Week 6**<br>**n = 7** |  |
| --- | --- | --- | --- | --- | --- | --- | --- | --- | --- | --- | --- | --- |
| **Age (years)** | 36 (29 – 51) |  | 35 (32 – 41) |  | 35 (29 – 51) |  | 36 (28 – 42) |  | 40 (27 – 51) |  | 29 (25 – 51) |  |
| **Sex (male: female)** | 2:6 |  | 3:4 |  | 1:6 |  | 2:4 |  | 4:3 |  | 3:4 |  |
| **Height (cm)** | 166 (160 – 175) |  | 178 (163 – 179) |  | 167 (162 – 176) |  | 174 (162 – 179) |  | 169 (165 – 178) |  | 178 (168 – 183) |  |
| **Weight (kg)** | 62 (60 – 67) |  | 78 (61 – 87) |  | 61 (57 – 70) |  | 63 (58 – 71) |  | 66 (62 – 75) |  | 64 (61 – 88) |  |
| **Body mass index (kg/m<sup>2</sup>)** | 21.5 (20.2 – 24.8) |  | 26.2 (22.1 – 28.1) |  | 22.2 (20.6 – 25.1) |  | 22.2 (20.0 – 24.5) |  | 22.6 (22.2 – 24.2) |  | 20.0 (19.7 – 26.3) |  |
| **Hypertension, n (%)** | 2 (25%) |  | 1 (14%) |  | 1 (14%) |  | 0 (0%) |  | 0 (0%) |  | 0 (0%) |  |
| **Diabetes mellitus, n (%)** | 0 (0%) |  | 0 (0%) |  | 0 (0%) |  | 0 (0%) |  | 0 (0%) |  | 0 (0%) |  |
| **Hyperlipidemia, n (%)** | 0 (0%) |  | 1 (14%) |  | 1 (14%) |  | 1 (17%) |  | 0 (0%) |  | 1 (14%) |  |
| **Smoking history, n (%)** | 0 (0%) |  | 0 (0%) |  | 0 (0%) |  | 0 (0%) |  | 0 (0%) |  | 0 (0%) |  |
|  | **Visit 1** | **Visit 2** | **Visit 1** | **Visit 2** | **Visit 1** | **Visit 2** | **Visit 1** | **Visit 2** | **Visit 1** | **Visit 2** | **Visit 1** | **Visit 2** |
| **Systolic blood pressure (mmHg)** | 118 (110 – 133) | 118 (111 – 127) | 115 (94 – 126) | 116 (95 – 123) | 114 (108 – 124) | 115 (105 – 124) | 112 (105 – 115) | 115 (108 – 120) | 116 (114 – 131) | 121 (107 – 131) | 116 (108 – 122) | 118 (105 – 120) |
| **Diastolic blood pressure (mmHg)** | 66 (65 – 86) | 71 (68 – 79) | 61 (58 – 71) | 64 (55 – 73) | 73 (64 – 84) | 70 (66 – 81) | 69 (64 – 72) | 68 (65 – 71) | 70 (69 – 75) | 66 (64 – 69) | 66 (54 – 69) | 68 (67 – 73) |

Data are expressed as n (%) or medians (interquartile ranges).

Major output parameters of FMD measurements are shown week by week in **Table 2**. There was an increasing trend in the measured FMD median values across the time period, reflecting the ability of the operator to capture the flow-mediated dilatation more accurately.

**Table 2.** Flow-mediated dilatation measurement parameters during the study.

|  | **Week 1**<br>**n = 8** |  | **Week 2**<br>**n = 7** |  | **Week 3**<br>**n = 7** |  | **Week 4**<br>**n = 6** |  | **Week 5**<br>**n = 7** |  | **Week 6**<br>**n = 7** |  |
| --- | --- | --- | --- | --- | --- | --- | --- | --- | --- | --- | --- | --- |
|  | **Visit 1** | **Visit 2** | **Visit 1** | **Visit 2** | **Visit 1** | **Visit 2** | **Visit 1** | **Visit 2** | **Visit 1** | **Visit 2** | **Visit 1** | **Visit 2** |
| **Baseline diameter (mm)** | 3.14 (2.77 – 4.02) | 3.32 (2.75 – 3.91) | 3.47 (2.90 – 4.06) | 3.51 (3.11 – 3.97) | 3.22 (2.55 – 4.29) | 3.12 (2.72 – 4.24) | 3.08 (2.70 – 3.98) | 2.95 (2.82 – 3.84) | 3.48 (3.04 – 3.99) | 3.50 (3.07 – 4.10) | 3.67 (2.95 – 4.53) | 3.55 (2.93 – 4.57) |
| **Flow-mediated dilatation (%)** | 2.95 (-0.12 – 6.85) | 3.68 (-0.95 – 6.34) | 2.82 (1.55 – 4.96) | 2.62 (1.12 – 3.04) | 5.44 (1.03 – 6.77) | 4.99 (0.68 – 8.43) | 5.69 (1.94 – 7.29) | 5.08 (1.91 – 7.44) | 2.37 (1.00 – 5.60) | 2.72 (0.77 – 5.84) | 5.05 (3.39 – 6.48) | 5.16 (4.42 – 7.43) |
| **Shear rate – baseline (s<sup>-1</sup>)** | 210 (133 – 254) | 168 (99 – 225) | 180 (122 – 207) | 171 (133 – 204) | 224 (138 – 266) | 176 (122 – 240) | 209 (151 – 266) | 208 (190 – 225) | 153 (107 – 243) | 159 (118 – 244) | 213 (181 – 222) | 218 (144 – 295) |
| **Shear rate – maximum (s<sup>-1</sup>)** | 442 (384 – 701) | 446 (276 – 677) | 512 (453 – 682) | 886 (370 – 1067) | 713 (574 – 786) | 760 (367 – 802) | 1051 (497 – 3136) | 862 (608 – 1002) | 744 (440 – 855) | 668 (420 – 847) | 1034 (649 – 1054) | 824 (504 – 1048) |

Data are expressed as medians (interquartile ranges).

Figure 1 shows the CV values for D and FMD. Both showed a decreasing trend over the training period. At week 6, both CV values were within the required range recommended by the recent guideline: below 2% for D and below 15% for FMD.^1^ The CV values at week 6 were 1.73% for D and 14.74% for FMD.

**Figure 1.**
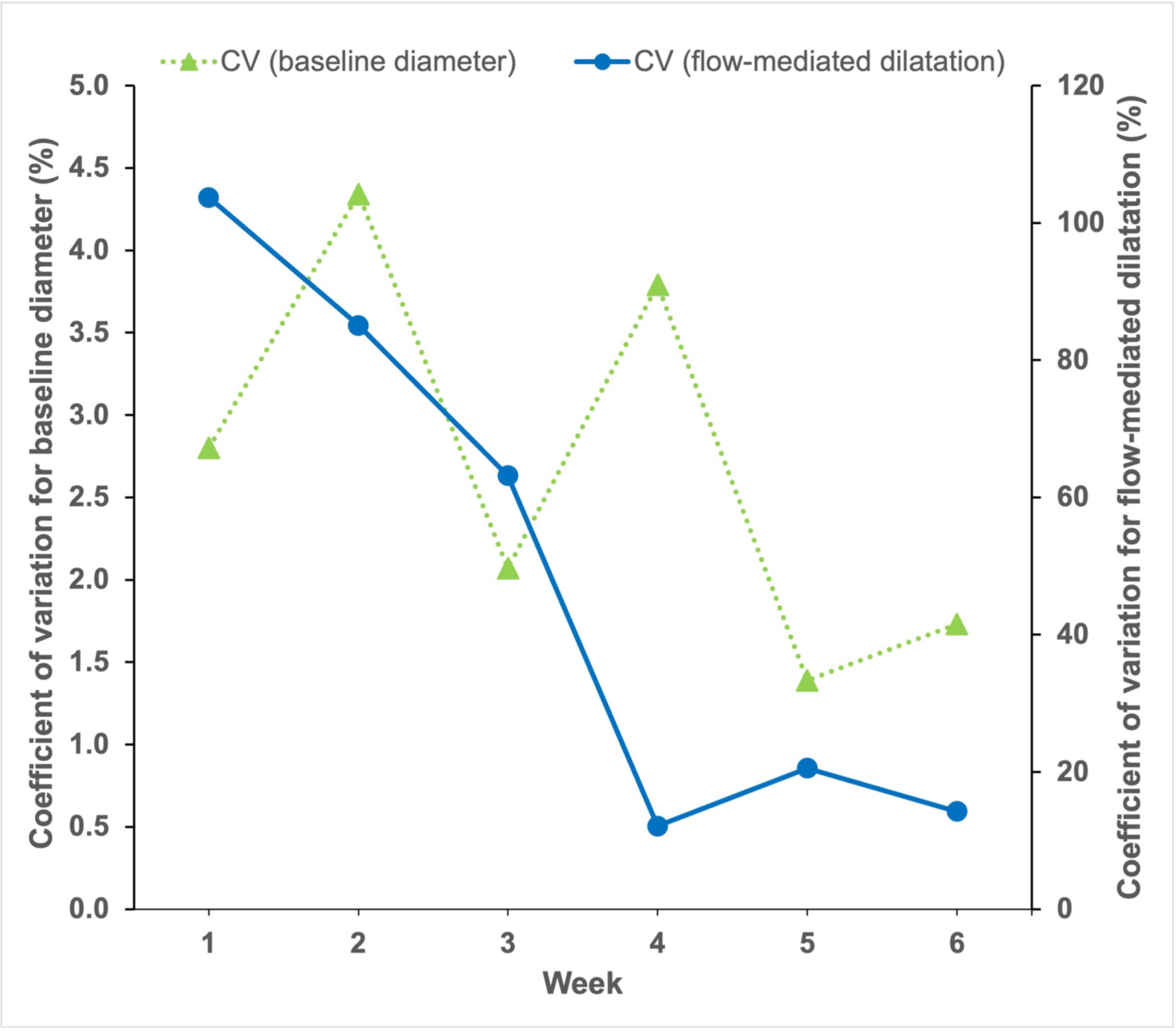
Changes in intersession coefficient of variation (CV) for baseline diameter and flow-mediated dilatation over time.

## Discussion

FMD is a valuable tool for the assessment of endothelial function, but multiple factors may hinder the attainment of reproducible results. One crucial factor is operator training, and as yet, no study has established the feasibility and duration of a proper training regimen for novices – as has been done with other techniques.^8, 10, 11^ Our study demonstrates a training program and the learning curve of a novice operator that culminated in the attainment of sound reproducibility between FMD measurements.

Different factors can influence measured FMD values and their reproducibility, including individual differences in arterial diameter and stiffness, as well as the amount of shear stress generated, diet, exercise, stimulant drink consumption and smoking history.^1, 2^ To minimize the influence of the aforementioned factors, we followed the recommendations outlined in the most recent guideline about FMD measurement.^1^ Regarding the training of operators, certain studies up to this point have mentioned training programs involving experienced sonographers, with benchmarks to assess sonographer proficiency. The training for sonographers with previous experience with FMD studies by Ghiadoni et al consisted of 20 supervised scans, and afterwards, quality certification was received when 5 consecutive scans were accepted by a core reading lab.^12^ Thijssen et al recommended that the training is sufficient when the inter-session CV for D is below 2% and the inter-session CV for FMD is below 15%.^1^ Since no emphasis has been placed on training of complete novices, our goal was to follow the learning curve of a complete novice under standardized circumstances, and describe a potential program for future training.

The training regimen outlined in this study was achieved within a reasonable 7-week time-frame, which comprised 6 weeks of measurements on various subjects – in addition to an earlier week of gaining basic procedural and theoretical understanding of the technique. The CV for both D and FMD showed a decreasing trend from the onset of the training period to the end, reinforcing that proper training can lead to sound reproducibility. The inter-session CV values of the last week for both D and FMD were below the threshold recommended in the recent FMD guideline.^1^

The CV values for D were below the recommended threshold at week 5 and 6. Beyond the gained experience and improved hand-eye coordination, precise placement of both the pressure cuff and ultrasound probe could play an important role in the attainment of these CV values. It has been established that placement of the pressure cuff can have a significant impact on FMD, mostly due to the potential differences in the volume of ischemic area distal to the cuff and consequential differences in the shear stress stimulus after cuff deflation.^13^ In this study, the distances of both the pressure cuff and ultrasound probe relative to the elbow crease were measured at the conclusion of the first visit, using a removable, skin-friendly marker and a ruler. The cuff was always placed distal to the elbow crease around the forearm of the subject. The key distance measurements were remade at the beginning of the second visit, allowing for a targeted placement of the probe and the cuff. The angle of transducer position was also recorded at the end of the first visit using a protractor, and this was followed in the subsequent visit as well. Targeted placement of the cuff and the probe were found to be helpful towards the achievement of reproducible results.

The CV for the FMD value also followed a decreasing trend over the study. The initial CV values for FMD were higher, and the FMD values were lower in the beginning of the study compared to the values obtained during the final week. This reflects the complexity of obtaining reliable FMD values, especially given how slight movements over the course of the measurement can markedly alter the results. The FMD values were initially low, suggesting that the sonographer was not able to capture the true FMD in subjects. Eventually, the CV values for FMD were within the recommended range at week 6, and the FMD values were higher compared to the beginning of the study, and reached the median (IQR) values of [5.05% (3.39% – 6.48%); Visit 1] and [5.16% (4.42% – 7.43%); Visit 2] by the final week – demonstrating that the operator could capture the diameter changes of the brachial artery more precisely. These FMD values measured at week 6 are physiologically reasonable based on the refence intervals published recently.^2^

It should be noted that this study has limitations. Firstly, it is based on the experience of a single novice operator, and may not capture the capabilities of others training in vascular measurement techniques. Second, the subject sample sizes for each week of measurement were small, ranging from 6 to 8 subjects per week. Third, for female subjects, the measurements were not performed at a standardized part of the menstrual cycle. Although the menstrual cycle has been found to impact endothelial function,^14^ since the time interval between the repeated measurements ranged between 1 and 3 days (with the median value of 1 day), we believe that this limitation did not have a major effect on our results. Despite these limitations, this study reflects a robust training regimen for FMD measurement that led to optimal reproducibility values by its conclusion.

Given that theoretical learning, together with practical skill acquisition were able to take place within 7 weeks and yield sound reproducibility values, our study suggests that standardized systems for learning FMD measurement can be implemented successfully and conveyed to inexperienced operators.

## Data Availability

All data produced in the present study are available upon reasonable request to the authors.

## Acknowledgements

This research was supported by the NIHR Cambridge Biomedical Research Centre (BRC-1215-20014). The views expressed are those of the authors and not necessarily those of the NIHR or the Department of Health and Social Care. This research was also supported by the Evelyn Trust Grant 20/58.

